# Development of at-home sample collection logistics for large scale SARS-CoV-2 seroprevalence studies

**DOI:** 10.1101/2021.01.14.21249824

**Authors:** Aishani V. Aatresh, Kate Cummings, Hilary Gerstein, Christopher S. Knight, Andreas Limberopolous, Megan A. Stasi, Alice Bedugnis, Kenneth A. Somberg, Camila T. França, Michael J. Mina

## Abstract

In the midst of a pandemic, serologic studies are a valuable tool to understand the course of the outbreak and guide public health and general pandemic management. However, given significant safety constraints including social distancing and stay-at-home orders, sample collection becomes more difficult given traditional phlebotomy protocols. For such studies, a representative sample of the underlying population is paramount to elicit meaningful insights that capture the spread of the infection, particularly when different sub-populations face varying disease burden. We aimed to address these challenges by conducting a fully remote study to investigate the seroprevalence of SARS-CoV-2 in the state of Massachusetts. Leveraging electronic study engagement and at-home self-collection of finger-prick samples, we enrolled 2,066 participants representative of the ethnic and racial composition of Massachusetts. SARS-CoV-2 total IgG seropositivity was 3.15%, and follow-up measurements at days 7, 15, 45, and 90 indicate a generally durable antibody response. A higher risk of infection was observed for healthcare workers and their cohabitants and those with comorbidities, as well as lower-income, less educated, Hispanic, and those in the age groups of 18-29 and 50-59-years-old. High engagement and positive feedback from the participants and quality of self-collected specimens point to the usefulness of this design for future population-level serological studies that more effectively and safely reach a broad representative cohort, thus yielding more comprehensive insights into the burden of infection and disease in populations.

**Key points:** *Question:* We aimed to implement a fully remote seroprevalence study for SARS-CoV-2, leveraging electronic methods and at-home self-collection of specimens to engage a representative study population.

*Findings:* The population enrolled reflected the ethnic and racial composition of Massachusetts, revealing a SARS-CoV-2 seroprevalence of 3.15% and higher risk of previous infection associated with healthcare workers/their cohabitants, those with comorbidities, lower-income, less educated, Hispanic, and those in age groups 18-29 and 50-59 years old.

*Meaning:* High engagement and positive feedback from participants as well as quality of self-collected specimens point to the usefulness of this design for future population-level serological studies.

## Introduction

The coronavirus disease 2019 (COVID-19) pandemic caused by severe acute respiratory syndrome coronavirus 2 (SARS-CoV-2) has had far-reaching consequences since its emergence in Wuhan, China, in December 2019^1^. As of mid-November, there have been over 55 million cases and 1.34 million deaths worldwide. The more subtle cost exacted upon society has been evident in the rise of virtual school, remote work, severe job loss, and economic contraction^2^.

Coronaviruses (*Coronaviridae*) are a family of enveloped positive-sense single-stranded RNA viruses with 39 species that infect vertebrates and have been responsible for causing three major epidemics over the past two decades through zoonotic spillover^3^— Severe acute respiratory syndrome (SARS) in 2002 caused by SARS-CoV, Middle East respiratory syndrome (MERS) in 2012 caused by MERS-CoV, and now COVID-19 caused by SARS-CoV-2^4^. These three viruses belong to the genus *Betacoronavirus*, which also includes the common cold-causing OC43 and HKU1 viruses. The binding-mediating spike proteins of SARS and SARS-CoV-2 share 76% homology at the amino acid level with 74% sequence identity of the receptor binding domain (RBD) with a shared mechanism of the angiotensin-converting enzyme 2 (ACE2) as the host cell receptor target for infection, confirmed by crystal structures^5^.

Testing and contact tracing both symptomatic and asymptomatic infected individuals is essential to characterize infection dynamics and control the spread of the current COVID-19 pandemic^6, 7^. Currently, reverse-transcriptase polymerase-chain-reaction tests (RT-PCR) form the bedrock of SARS-CoV-2 testing in the United States, detecting copies of viral RNA in an infected patient. This strategy, however, has been limited by supply chain breakdowns, lack of personal protective equipment (PPE) and medical supplies^8^, testing backlogs, reliance on trained personnel^9^, and false positives from significantly amplified but non-infectious viral loads^10^. Alternatively, the measurement of antibodies against SARS-CoV-2 in blood is relatively cheap, and serology has been proposed as an alternative method to identify individuals who have previously had symptomatic or asymptomatic SARS-CoV-2 infections and recovered^11^.

Studies surrounding the humoral response mounted against SARS-CoV-2 infection continue to emerge as the pandemic persists^12, 13, 14^. Current research demonstrates seroconversion for IgA, IgM, and IgG antibodies over the course of infection, but it is still unclear how long-lived and functional (e.g. potently neutralizing) these responses are after the primary infection. Although our understanding of the clinical significance of seropositivity is still limited, well designed population sero-surveys can be powerful tools to help determine the stage of the outbreak and trend of disease^15^. Such studies will also provide a better understanding of the dynamics of antibody responses for differentiation of individuals with acquired immunity from those who remain susceptible to infection and disease, therefore helping to determine how to safely reopen the economy, where to deploy resources for disease prevention and management, and will help identify emerging outbreaks early^16^.

In order to facilitate the use of serology as a public health tool, we designed and implemented a fully remote mechanism for conducting large-scale sero-surveys. We coupled the use of electronic medium for study engagement and successful recruitment and retention of representative cohorts with at-home self-collection of serological specimens, allowing for greater participant safety and increased sampling of diverse populations with reduced cost. Using these logistics, we successfully conducted a cross-sectional survey of the population of Massachusetts, measuring the prevalence of total IgG antibodies to SARS-CoV-2 in symptomatic and asymptomatic individuals using simple at-home finger prick samples. Findings may be used to inform evolving health policy in response to the pandemic and provide a proof-of-concept for the logistics of future sero-epidemiological studies.

## Materials and Methods

### Study Design

#### Recruitment

This at-home, decentralized SARS-CoV-2 seroprevalence study targeted adult (≥ 18 years of age) residents of Massachusetts, with no requirements around prior or expected exposure to SARS-CoV-2. With the goal of enrolling approximately 2,000 volunteers, potential participants were identified through partnerships with for- and non-profit entities and digital ad campaigns and referrals, and they received a link to a landing page to learn more about the study and enroll if interested. Participants were required to have reliable Internet access. Although not included as a formal exclusion criterion, participants were also required to speak English, as the study was not offered in additional languages. If eligible, based on electronic screening, participants electronically reviewed the informed consent form and completed a background questionnaire (Supplementary Table 1) with questions related to demographic profile (including sex, age, race, ethnicity, region of residency, education, income range, housing status, pregnancy and recent medical history/comorbid conditions), and COVID-19 history (including presumptive and confirmed SARS-CoV-2, checklist of symptoms and their duration, level of care received and clinical outcome). Socio-behavioral questions related to perception of COVID-19 risk (such as adherence to social distancing guidelines, use of masks/face coverings in public and type of transportation used) were also included. Volunteers were not compensated for their participation in the study, and there were no requirements regarding prior or expected exposure to SARS-CoV-2.

**Table 1:**
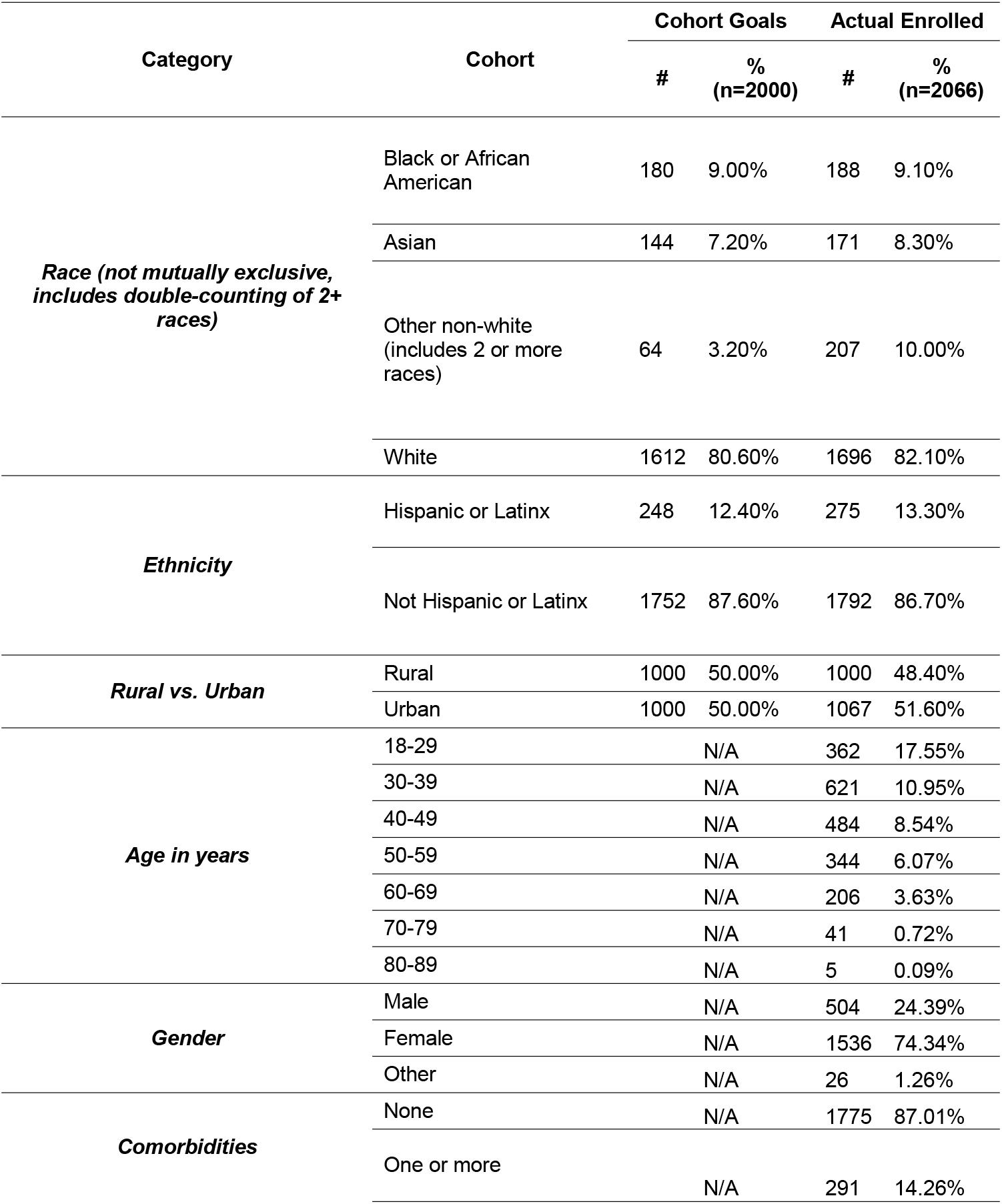
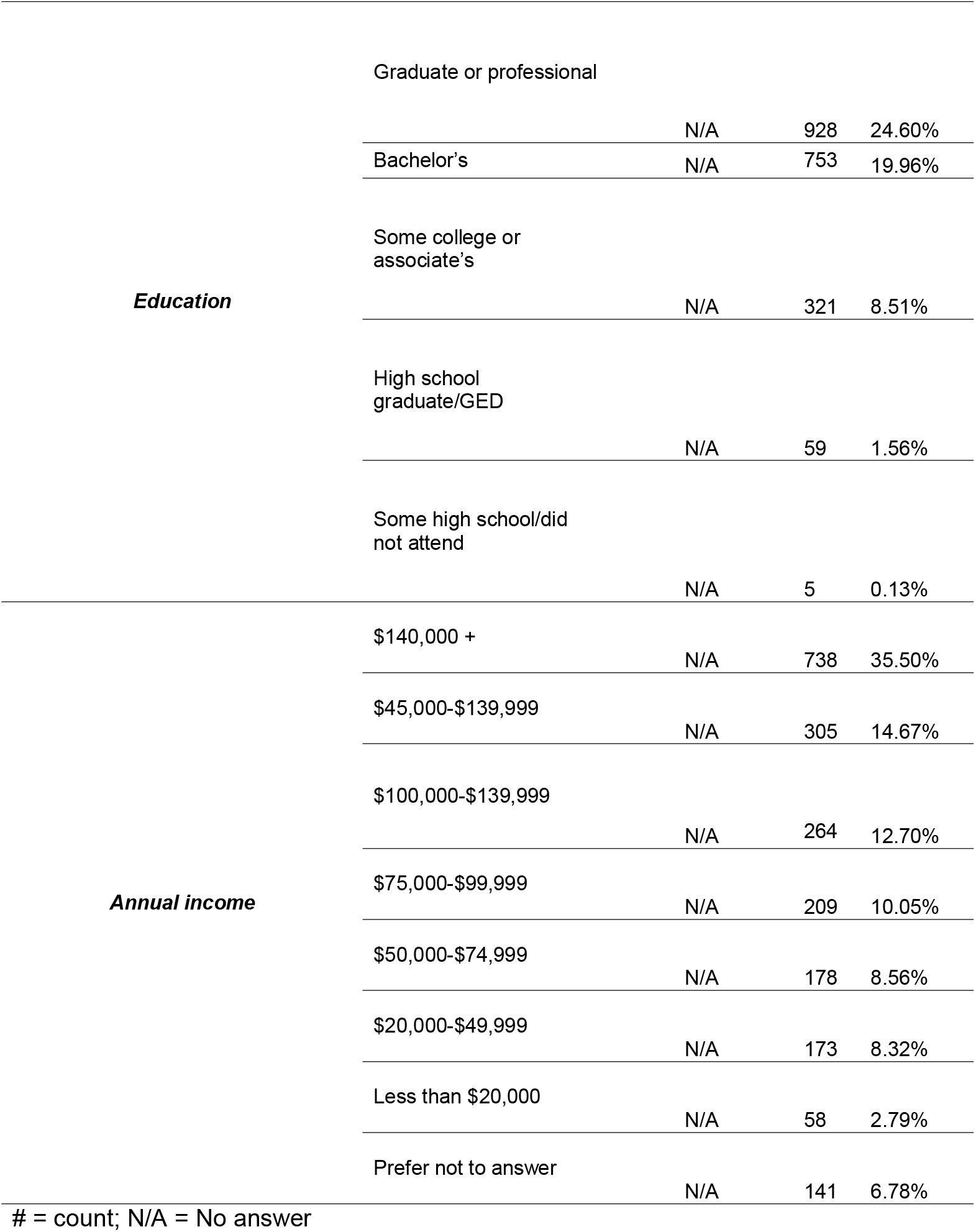
Demographic profile of study participants

#### Specimen collection

After completing the baseline questionnaire, participants were shipped through the United States Postal Service (USPS) an at-home specimen collection kit which included two spring-loaded lancets, a biohazard bag, and instructions for self-administered finger-prick blood collection. Participants were asked to place approximately 10-20 drops of blood on to the supplied Whatman 903 dried blood spot protein saver filter paper. After air drying the specimen, the participants were instructed to place the filter paper into sealed, pre-paid envelopes provided in the kit and mail it to the central laboratory, Molecular Testing Labs, a CLIA-licensed and CAP-accredited laboratory, for analysis. All participants with a positive SARS-CoV-2 IgG result were asked to provide additional blood finger-prick samples at day 7, 15, 45, and 90 after receiving the initial result (Figure 1a and b). Throughout the study, all participants had access to frequently asked questions (FAQ) as well as a dedicated support team online or by phone to discuss questions about specimen collection or other elements of the study.

**Figure 1:**
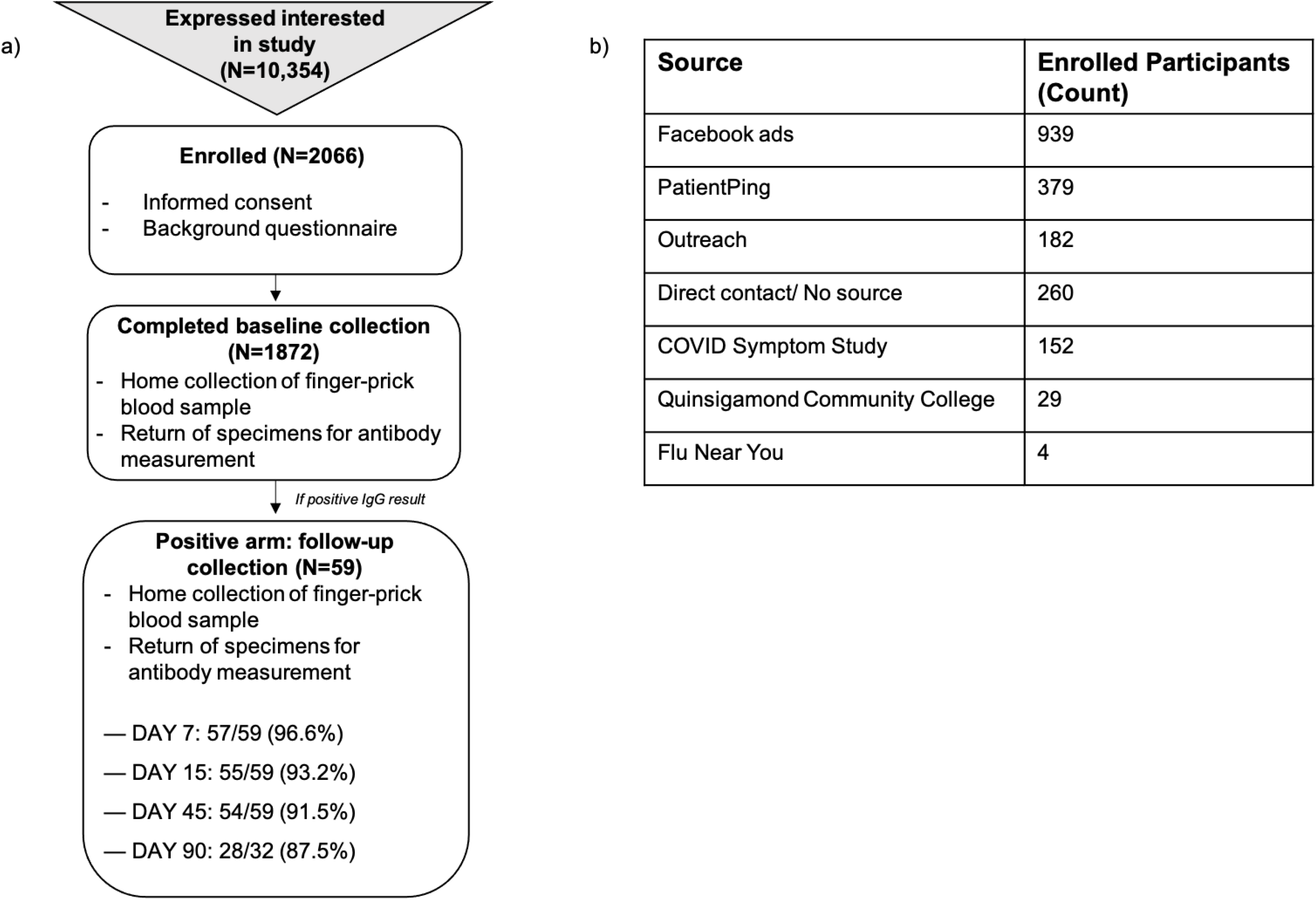
Study design. a) Progression of study from recruitment and participant admission to testing and follow-up sample collection for individuals who test positive for IgG antibodies against SARS-CoV-2 at baseline. b) Counts of study population garnered through each method; b) Known sources and counts of participants recruited electronically across the state of Massachusetts.

#### Laboratory tests

The presence of total IgG antibodies against the S1 protein of SARS-CoV-2 was measured using the FDA EUA-approved EUROIMMUN ELISA assay as previously described^17^. Test results were returned to the participants by Molecular Testing Labs within 24-72 hours of receipt of the specimen using the study mobile application platform as positive, negative, or indeterminate. A second kit was offered to any participant who received an indeterminate result and wished to provide another specimen.

### Statistical Analysis

Chi-square tests and univariate logistic regression models were used to investigate the association between demographic, clinical, and behavioral factors and seroprevalence of antibodies against SARS-CoV-2. All analyses were performed using Python (version 3.8.5).

### Ethics approval

Ethical clearance was obtained from Advarra (Pro00043729) and the Harvard T.H. Chan School of Public Health review board (IRB20-1511). Written informed consent was obtained electronically from all participants prior to enrollment in this study.

## Results

### Study Enrollment and Participant Demographics

690 of the planned 2,000 participants were enrolled between June 16 and June 30, 2020 using convenience sampling (Figure 1a) through partnerships leading to employee and symptom-tracking app referral (Figure 1b). Most of this initial population was comprised of Caucasian, high-income (>$140,000) individuals (Table 1). In order to increase diversity and mirror race and ethnicity proportions of 2019 Massachusetts census data and achieve a 50/50 split between residency within rural or urban centers (as defined by the Massachusetts State Office of Rural Health), age, zip code, internet access, race and ethnicity information were used to pre-screen interested individuals and temporarily place them on a waiting list/lottery, allowing for the random selection of individuals to fulfill the cohort population profile. The remaining participants (n=1,376) were enrolled between July 29 and August 24, 2020. In total, 48.3% (n=939) of participants for whom recruitment data was available (n=1,945) were recruited through online ads, specifically Facebook (Figure 1b).

From an initial cohort of 2,066 study participants, 90.61% (n=1872) of individuals mailed their sample to the laboratory for analysis (Figure 1a). Only data from individuals who completed sample collection and return was included in the analysis. The median age of study enrollees was 40 years old (interquartile range [IQR] 32 to 52 years old), 73.95% (n=1368) were female, while 81.37% (n=1681) hold an undergraduate degree or higher (Table 1). The cohort was generally distributed over the state of Massachusetts (Supplementary Figure 1), with 48.65% and 51.35% from rural and urban areas, respectively. A small proportion of the 59 participants who tested positive (8.47%, n=5) cohabitated with other individuals in the study, and 13.24% of participants (n=245) were either healthcare workers or shared a household with one. A total of 40.11% (n=742) reported having symptoms resembling those of COVID-19 since January 2020 (including cough, fever, shortness of breath, sore throat, and new loss of smell or taste) (Figure 3c), and 14.09% (n=291) reported having one or more comorbid health conditions known to increase risk of COVID-19 (e.g., diabetes, asthma, being immunocompromised, heart or lung disease) (Table 1).

**Figure 2:**
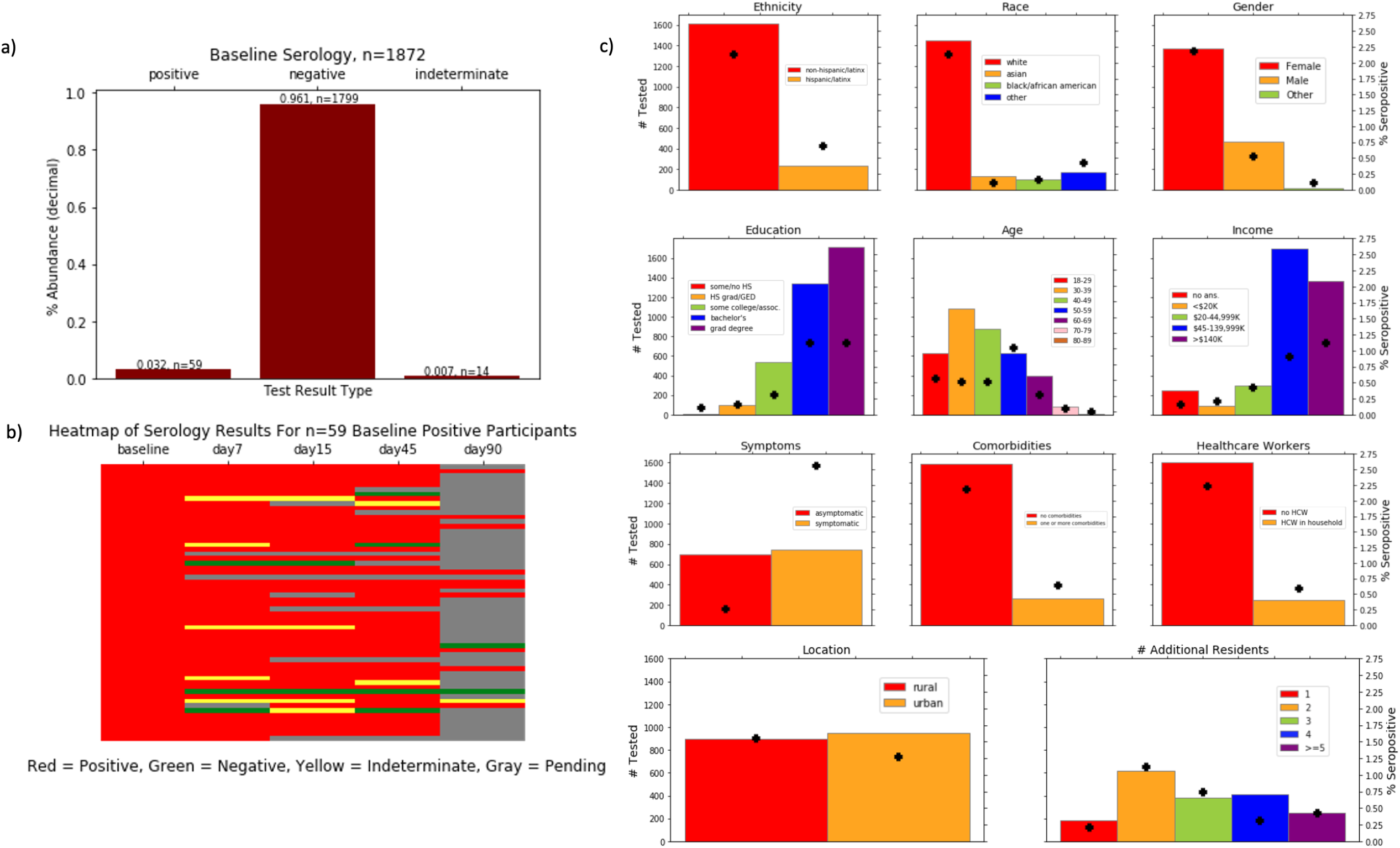
Seroprevalence of SARS-CoV-2 total IgG antibodies in Massachusetts. a) Distribution of positive, negative and indeterminate results for presence of total IgG antibodies against SARS-CoV-2 S1 protein across all individuals who returned a baseline test specimen (n=1872). b) Heatmap showing presence of total IgG antibodies against SARS-CoV-2 S1 protein in follow-up samples of individuals who tested positive at baseline. Each row represents an individual and each column a time-point of sample collection (baseline, days 7, 15, 45 and 90) with data as of November 17, 2020. c) Histograms showing the total counts (left y-axis) for each variable in the study population. Black crosses represent percentage seropositivity (right y-axis) against the entire population (n=1872) given individuals for each group.

**Figure 3:**
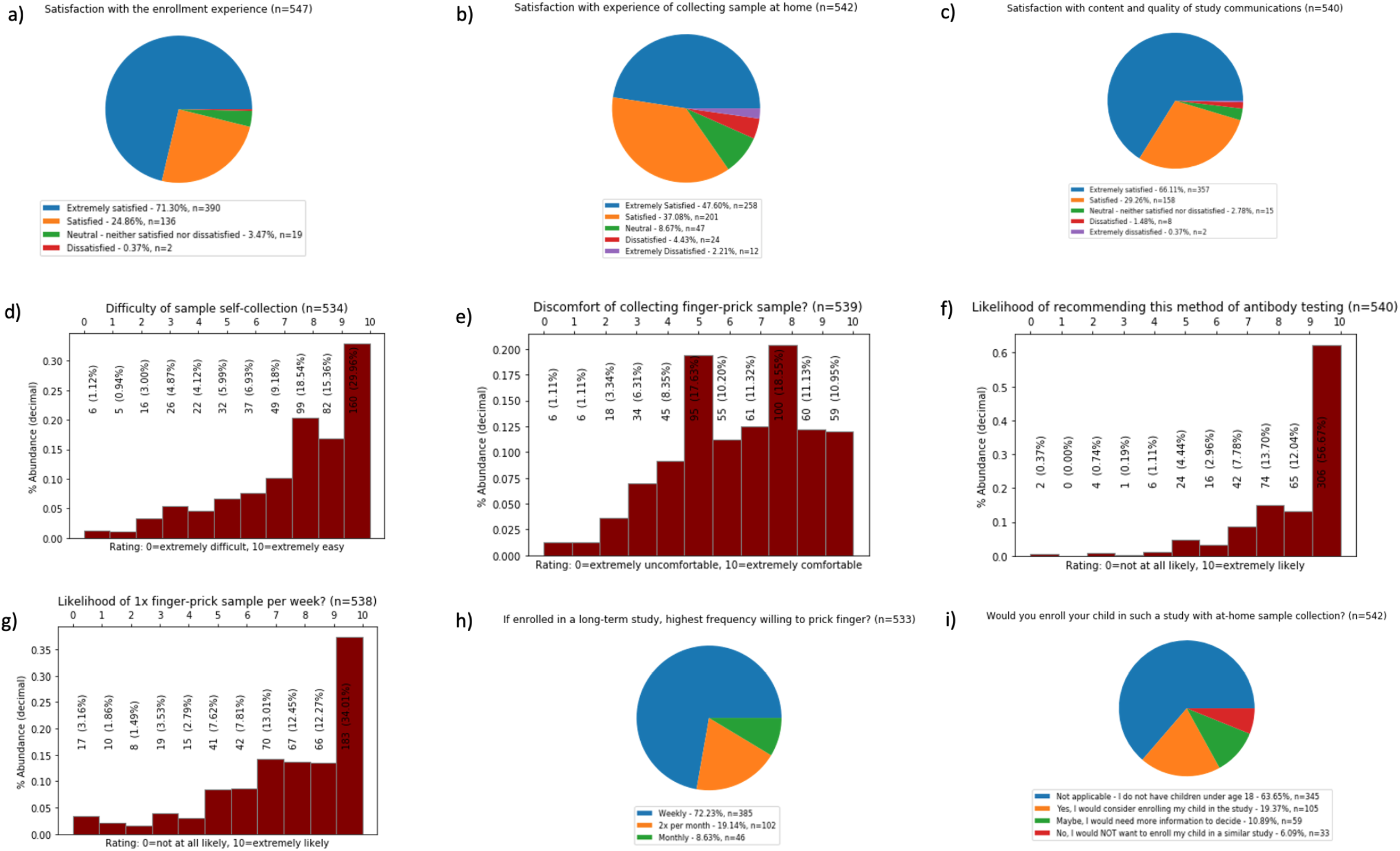
Participant feedback. Distribution of the participants who responded the feedback survey by satisfaction with study participation (a, b, c), difficulty with self-collection of blood sample (d, e), and willingness to collect samples at higher frequency, recommend this study to others or enroll their child(ren) in similar studies (f, g, h, i).

### SARS-CoV-2 serology in Massachusetts

3.15% (n=59) of the individuals who returned their samples were seropositive for total IgG antibodies against SARS-CoV-2 S1 protein (Figure 2a) and were requested to send follow-up samples at days 7, 14, 45, and 90 after initial positive result (Figure 2b). 31.25% (n=10 of 32) of these baseline seropositive participants for whom data is currently available, with follow-up ongoing, remained positive during the entire 90-day follow-up period, with 1 seemingly false positive at baseline and 5 indeterminate cases (Figure 2b).

### Risk of SARS-CoV-2 in Massachusetts

Chi-square goodness-of-fit tests and log odds scores (with base *e*) were used to assess the effect of different demographic or self-reported variables on seropositivity as an indication for past infection (Table 2). In this cohort, the individuals in the youngest age group (18-29 years old) and those 50-59 years old were at the highest risk of infection (log odds=0.137 and 0.805 respectively, *p*=*0*.*0157*). Hispanics had higher risk compared to non-Hispanic individuals (log odds=0.825, vs. log odds= −0.825, *p*=*0*.*0173*). Individuals with some or no high school were also shown to be at higher risk compared to any other educational level (log odds=3.155, *p*=*0*.*000014*), as well as those in the lowest income bracket, less than $20,000 in annual income (log odds=1.137, *p*=*0*.*0252*), and those who presented with any symptoms (log odds=1.137, *p*=*0*.*252*). Individuals sharing a household with others were generally at higher risk than those living alone (log odds= −0.25 to 0.23, p<0.001), although there were very small sample size effects in this analysis, rendering such conclusions extremely limited (Table 2).

**Table 2:** Association between demographic variables of interest and seropositivity

| Category | Cohort | # Seropositive/ # Tested | Log Odds Ratio (95% CI) |
| --- | --- | --- | --- |
| **Ethnicity (p=0.0173)** | Non-Hispanic/Latinx | 40/1613 | -0.825 (-1.47, -0.18) |
|  | Hispanic/Latinx | 13/237 | 0.825 (0.18, 1.47) |
| **Race\* (p=0.421)** | White | 40/1449 | -0.166 (-0.80, 0.47) |
|  | Asian | 2/128 | -0.747 (-2.25, 0.76) |
|  | Black/African American | 3/101 | -0.09 (-1.37, 1.18) |
| **Gender (p=0.0557)** | Female | 41/1368 | 0.077 (-0.34, 0.49) |
|  | Male | 10/465 | -0.26 (-0.95, 0.42) |
| **Education (p=0.000014)** | Some/no high school | 43866 | 3.155 (1.34, 4.97) |
|  | High school graduate/GED | 3/49 | 0.830 (-0.37, 2.03) |
|  | Some college/associate's degree | 6/270 | -0.224 (-1.08, 0.63) |
|  | Bachelor's degree | 21/669 | 0.131 (-0.39, 0.64) |
|  | Graduate or professional degree | 21/857 | -0.124 (-0.64, 0.39) |
| **Age in years (p=0.0157)** | 18-29 | 10/313 | 0.137 (-0.56, 0.84) |
|  | 30-39 | 9/540 | -0.530 (-1.25, 0.20) |
|  | 40-49 | 9/439 | -0.319 (-1.04, 0.41) |
|  | 50-59 | 19/314 | 0.805 (0.25, 1.36) |
|  | 60-69 | 5/198 | -0.105 (-1.04, 0.83) |
|  | 70-79 | 1/41 | -0.141 (-2.15, 1.87) |
|  | 80-89 | 0/5 | 0 |
| **Annual income (p=0.0252)** | ≤ \$20,000 | 4/47 | 1.137 (0.075, 2.20) |
| | \$20,000-44,999 | 8/150 | 0.636 (-0.13, 1.40) |
| | \$45,000-139,999 | 17/848 | -0.377 (-0.93, 0.18) |
| | ≥ \$140K | 21/681 | 0.064 (-0.45, 0.58) |
| **Symptoms (p=0.000252)** | *Asymptomatic* | 5/699 | -2.262 (-3.19, -1.33) |
|  | *Symptomatic* | 48/742 | 2.262 (1.33, 3.19) |
| **Comorbidities (p=0.107)** | *No comorbidities* | 41/1589 | -0.599 (-1.26, 0.058) |
|  | *One or more comorbidities* | 12/261 | 0.599 (-0.058, 1.26) |
| **Healthcare Workers (p=0.152)** | *Not a healthcare worker/no healthcare workers in household* | 42/1605 | -0.559 (-0.91, -0.20) |
|  | *Healthcare worker/one or more healthcare workers in household* | 11/245 | 0.559 (-0.12, 1.24) |
| **Location (p=0.449)** | *Rural* | 29/900 | 0.250 (-0.30, 0.80) |
|  | *Urban* | 24/950 | -0.250 (-0.80, 0.30) |
| **# of Additional Residents in Household (p=0.0000140)** | 1 | 4/183 | -0.301 (-1.33, 0.72) |
|  | 2 | 21/616 | 0.153 (-0.37, 0.67) |
|  | 3 | 14/383 | 0.225 (-0.38, 0.83) |
|  | 4 | 6/415 | -0.725 (-1.58, 0.13) |
|  | ≥ 5 | 8/253 | 0.0753 (-0.68, 0.83) |

There was no significant difference between those living in rural or urban areas, those who were or share a house with a healthcare worker, or between those with or without comorbidities, although the low number of participants in some of these groups may have precluded our analysis (Table 2). Of note, mask wearing and social distancing were not included in our analysis because of the general uniformity of this self-reported data across the study population; all but one person reported wearing a mask always or sometimes when outside of their household; the same held for all but six people with social distancing.

### Participant Feedback

After sample collection and testing were finalized, 1,764 participants were sent a survey to provide feedback about the study process, eliciting a 31% (n=547) overall response rate. Survey respondents were generally representative of the study population (Supplementary Figure 2). 96.16% (n=526) of them reported being extremely satisfied or satisfied with the process of enrolling in the study (Figure 3a), 84.68% (n=459) reported being extremely satisfied or satisfied with the experience of self-collection of the finger-prick sample (Figure 3b), with the majority of responses indicating sample collection was very to extremely easy (Figure 3d). More respondents rated the experience as more comfortable than not (Figure 3e). Meanwhile, 95.37% (n=515) were extremely satisfied or satisfied with the content and quality of study communications (Figure 3c). With respect to the potential for future studies, 56.67% (n=306) of respondents said they were extremely likely to recommend this method of remote enrollment, at-home self-collection of specimens and antibody testing to others (Figure 3f). 72.23% (n=385) of the responders were willing to self-perform finger-prick blood collection up to once per week if needed (Figure 3g). While 63.65% (n=345) of the respondents did not have children, 19.37% (n=105) of those who did indicated that they would enroll their child in such a study.

## Discussion

The risk of viral transmission and limited capacity of healthcare systems called for the decentralized, at-home nature of this SARS-CoV-2 seroprevalence study, leveraging online recruitment, eConsent, electronic questionnaires, and direct-to-patient shipping to reach a broad representative study population. This study was a valuable opportunity to utilize and assess an at-home approach, and participant survey data reveals it was overwhelmingly well-received and indicates a strong likelihood of success for future deployment of larger studies of this modality. Although the discomfort of the finger-prick was the biggest concern expressed by participants, self-collection of samples was reported to be easy and generated samples of quality without the need for trained professionals or PPE, providing a remedy for the difficulties often encountered when obtaining standard specimens by phlebotomy, particularly during a pandemic. While representative cohorts are especially important for COVID-19 prevalence estimation because of the disproportionate impact of that this pandemic has exacted upon racial and ethnic minorities^18^, minimally biased data regarding the status of the pandemic has been significantly limited thus far^19, 20, 21^. Convenience sampling can skew data by drawing a study cohort that is not representative of the underlying population, as such surveys may not be able to adequately reach less advantaged communities, whether in rural areas or in lower-income urban settings. Furthermore, individuals seeking or willing to receive testing may be more likely to have experienced illness.

To this end, the recruitment strategy employed in the present study was very successful in reaching a representation of the population structure seen across Massachusetts in regard to race, ethnicity, and location of residency. However, recruitment was still subject to skew towards individuals who were more prolific Facebook users, female, highly educated, and wealthy (>$140,000 annual income). A small number of participants were not fully random because of shared households and thus could be non-independent exposures, possibly skewing any extrapolated seroprevalence estimates. Travel patterns were not accounted for, which may explain the similar risk observed among those from rural and urban areas. It is also possible that participants were generally more attuned to the nature of the pandemic and thus may have been less likely to have been infected because of greater behavioral and health awareness. Despite limitations, this study provides valuable data with regards to disease risk trends. Our data generally points to healthcare workers or those who live with them, those with comorbidities, and lower-income, less educated, and Latin-American individuals as those with relatively greater risk for COVID-19 in Massachusetts. Individuals in the age ranges of 18 to 29 and 50 to 59 years were more likely to have antibodies to SARS-CoV-2, which likely reflect behavioral patterns (e.g., less careful social behavior) and increased transmission among the young and a covarying increased risk of disease by greater incidence of comorbidities among the older, respectively. For the 59 of 1872 individuals with positive IgG to SARS-CoV-2 at baseline, sustained serological responses are generally observed, with the few negative results at day 90 possibly serving as an indicator of the natural waning of an antibody response over time^22^. Some of the samples may appear as a false negative or a false positive also because the test sensitivity and specificity, respectively, are predicted to be lower for samples of lower antibody prevalence^17^.

The population breakdown in Massachusetts does not accurately reflect the disease burden for COVID-19 reported in macro-aggregated statistics across the country, and while it is important to consider the limitations of our data when making claims about seroprevalence for the state, it also important to recognize the heterogeneity of the pandemic in the state and nationwide. At the time of this study in early fall of 2020, the incidence of COVID-19 was relatively low in the state of Massachusetts, and therefore follow-up studies in more disease-burdened areas and/or with larger sample sizes would provide the greater power needed to make more definitive assessments about risk for COVID-19 for specific demographic groups. The overall seroprevalence observed in this study (3.15%) indicates that the Massachusetts population is nowhere near close to supposed herd immunity^23^ and demonstrates the importance of a continued commitment to fighting the pandemic through controllable non-pharmaceutical interventions currently at our disposal. Future studies would improve recruitment of such a representative sample by taking into account income and education levels in conjunction with racial and ethnic data to more accurately represent complete socioeconomic conditions.

## Data Availability

Data has not been made publicly available to avoid compromising participant privacy or violating the ethical agreement in the informed consent forms. Data is available upon reasonable request by contacting Michael J. Mina.

## Acknowledgements

We are grateful to all the volunteers who participated in this study. We would like to thank the TrialSpark Clinical Research Coordinators and Recruitment, Operations, Data, Marketing and Technology teams for partnering on the development and successful deployment of the logistics used in this study. We thank Molecular Testing lab for IgG measurements. This study was funded by Open Research. M.J.M. is supported by the NIH Director’s Early Independence Award DP5OD028145.

## Abbreviations

ACE2: angiotensin-converting enzyme-2
CAP: College of American Pathologists
CLIA: Clinical Laboratory Improvement Amendments
COVID-19: coronavirus disease 2019
ELISA: enzyme-linked immunoassay
EUA: Emergency Use Authorization
FDA: Food and Drug Administration
IgG: immunoglobulin G
PPE: personal protective equipment
RBD: receptor binding domain
RT-PCR: reverse transcriptase polymerase chain reaction
SARS-CoV-2: severe acute respiratory syndrome coronavirus 2
SARS: severe acute respiratory syndrome
USPS: United States Postal Service

## Figure legends

*Supplementary table 1: Participant profile questionnaire*

Background questionnaire completed by participants after informed consent was obtained, gathering self-reported demographic and clinical data.

*Supplementary figure 1: Participant map*

Distribution of zip codes of residence for all study participants across the state of Massachusetts. Map created using Google My Maps.

*Supplementary figure 2: Representative survey sample*

Comparison of general distribution of survey sample (maroon, n=542) against general sample distribution of study population (green, n=2066/as data is available for age, n=2063) for the commonly collected demographic variables of income, ethnicity, and age.

