## Supplementary Table 1, Supplementary figure 1 and Supplementary figure 2 for "Development of at-home sample collection logistics for large scale SARS-CoV-2 seroprevalence studies"

### Supplementary table 1: Participant profile questionnaire

| Question | Possible Answers | Follow-up Questions and Notes |
|---|---|---|
| *Gender* | Female, Male, Transgender female, Transgender male, Other gender identity not listed, prefer not to answer | If female, pregnancy<br>If pregnant, which trimester |
| *Ethnicity* | Hispanic or Latinx, not Hispanic or Latinx | |
| *Race* | Black or African American, American Indian or Alaskan Native, Asian/Hawaiian/Pacific Islander, White, Other | multiple selection for individuals who are more than one race |
| *Transportation* | Own car, public transportation, ride shares/shared cars, none | |
| *Residence* | Apartment, condo, single-family home, other | If apartment/condo: # of units in building |
| *Highest level of education* | some high school or did not attend high school, high school graduate/GED, some college or associate degree, bachelor's degree, graduate or professional degree | |
| *Annual household income* | $140,000 or more, $100,000–$139,999, $75,000–$99,999, $50,000–$74,999, $20,000–$49,999, less than $20,000, prefer not to answer | options were modified partway through enrollment to better stratify $45,000–$139,999 |
| *Basic metrics* | Height (feet and inches), Weight (pounds) | |
| *Comorbidities* | Heart disease, immunocompromised or being treated for cancer/diabetes/kidney disease/dialysis, liver disease, chronic lung disease, moderate to severe asthma, smoking or vaping history | |
| *Number of people in household, whether anyone in it had been diagnosed with COVID-19, whether they or anyone else lives with a healthcare worker* | # people, Yes/No | |
| *Whether they have spent 3 minutes or more within 6 feet of someone with confirmed COVID-19 (i.e., were they exposed to COVID-19)* | Yes/No | Best estimate of the date that they were exposed |
| *Whether they think they may have been infected with COVID-19 at some point since February 2020* | Yes/No | If they may have been infected, COVID-19 symptoms experienced: fever, cough, shortness of breath or difficulty breathing, chills, repeated shaking with chills, muscle pain, headache, sore throat, new loss of smell or taste, no symptoms); best estimate of the date that the symptoms began; estimated duration of the symptoms (1-2 days, 3-7 days, more than 7 days, symptoms are ongoing |
| *Whether they sought treatment for COVID-19* | Yes/No | If yes, level of treatment received: visit with a healthcare professional via phone/video or office visit, urgent care or emergency room visit, home care visit, admitted to the hospital, admitted to the intensive care unit, was not able to get treatment, whether they were put o a ventilator |
| Whether they were tested for the novel coronavirus that causes COVID-19 AND Whether they had a blood test to test for antibodies to the novel coronavirus that causes COVID-19 | Yes/No | If yes: how they were tested (at home, doctor's office, laboratory company, hospital, drive-through, other), when they were tested, the results of the test, if known AND whether the antibody test result was positive or negative |
| *Whether they wear a mask or face covering when out in public AND Whether they practice social distancing when out in public, defined as maintaining a distance from other people of at least 6 feet* | Yes/Sometimes/No | |

**Supplementary figure 1: Participant map**

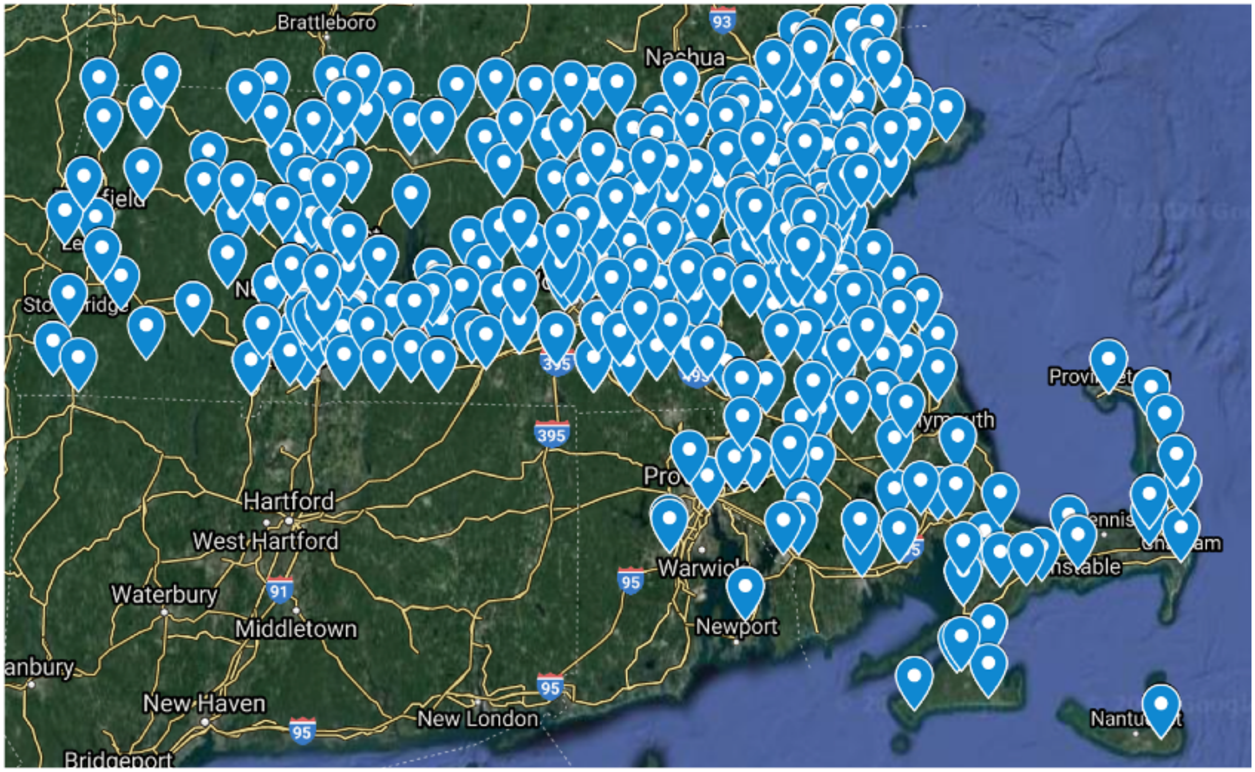

### Supplementary figure 2: Representative survey sample

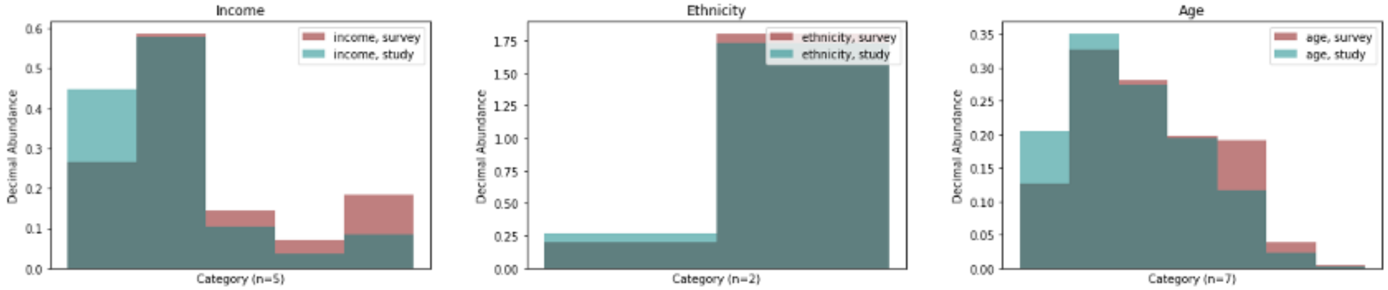
